## Supplementary Material for "Associations of age at diagnosis of breast cancer with incident myocardial infarction and heart failure: a prospective cohort study"

**Table S1.** Ascertainment of breast cancer and age at breast cancer diagnosis.

**Table S2.** Ascertainment of myocardial infarction and heart failure.

**Table S3.** Definition and assessment of covariates.

**Table S4.** Baseline characteristics of participants by breast cancer status after propensity score matching (n=64 964).

**Table S5.** Associations of age at breast cancer diagnosis with incident myocardial infarction and heart failure among participants with breast cancer: competing risk models (n=16 241).

**Table S6.** Associations of breast cancer with incident myocardial infarction and heart failure among different diagnosis age groups after propensity score matching: competing risk models (n= 64 964).

**Table S7.** Associations of age at breast cancer diagnosis with incident myocardial infarction and heart failure among participants with breast cancer after excluding myocardial infarction and heart failure diagnosed within 5 years since baseline (n=15 589).

**Table S8.** Associations of breast cancer with incident myocardial infarction and heart failure among different diagnosis age groups after excluding myocardial infarction and heart failure diagnosed within 5 years since baseline, results from propensity score matching analyses (n=62 356).

**Table S9.** Associations of age at breast cancer diagnosis with incident myocardial infarction and heart failure among participants with breast cancer after excluding participants aged <50 years at baseline (n=14 000).

**Table S10.** Associations of breast cancer with incident myocardial infarction and heart failure among different diagnosis age groups after excluding participants aged <50 years at baseline, results from propensity score matching analyses (n=56 000).

**Table S11.** Associations of age at breast cancer diagnosis with incident myocardial infarction and heart failure among participants with breast cancer when the follow-up period ends on December 31, 2019 (n=15 909).

**Table S12.** Associations of breast cancer with incident myocardial infarction and heart failure among different diagnosis age groups when the follow-up period ends on December 31, 2019, results from propensity score matching analyses (n=63 636).

**Table S13.** Associations of age at breast cancer diagnosis with incident myocardial infarction and heart failure among participants with breast cancer after further adjusting for menopausal status, breast cancer surgery, and hormone replacement therapy (n=16 241).

**Table S14.** Associations of breast cancer with incident myocardial infarction and heart failure among different diagnosis age groups after further adjusting for menopausal status, breast cancer surgery, and hormone replacement therapy, results from propensity score matching analyses (n=64 964).

**Table S15.** Comparison of baseline characteristics between participants included (n=251 277) and excluded due to history of myocardial infarction or heart failure, without complete data on low-density lipoprotein cholesterol, or having myocardial infarction or heart failure before breast cancer at follow-ups (n=22 048).

**Figure S1.** Subgroup analyses to identify potential modifying effects from covariates on the associations between breast cancer and incident myocardial infarction in participants with breast cancer diagnosed at age <50 and their controls by using Cox proportional hazards models (n=10 696).

**Figure S2.** Subgroup analyses to identify potential modifying effects from covariates on the associations between breast cancer and incident myocardial infarction in participants with breast cancer diagnosed at age 50 to 59 and their controls by using Cox proportional hazards models (n=22 548).

**Figure S3.** Subgroup analyses to identify potential modifying effects from covariates on the associations between breast cancer and incident myocardial infarction in participants with breast cancer diagnosed at age ≥60 and their controls by using Cox proportional hazards models (n=31 720).

**Figure S4.** Subgroup analyses to identify potential modifying effects from covariates on the associations between breast cancer and incident heart failure in participants with breast cancer diagnosed at age <50 and their controls by using Cox proportional hazards models (n=10 696).

**Figure S5.** Subgroup analyses to identify potential modifying effects from covariates on the associations between breast cancer and incident heart failure in participants with breast cancer diagnosed at age 50 to 59 and their controls by using Cox proportional hazards models (n=22 548).

**Figure S6.** Subgroup analyses to identify potential modifying effects from covariates on the associations between breast cancer and incident heart failure in participants with breast cancer diagnosed at age ≥60 and their controls by using Cox proportional hazards models (n=31 720).

**Figure S7.** Cubic spline curves of the association between diagnosis age of breast cancer and incident myocardial infarction.

**Figure S8.** Cubic spline curves of the association between diagnosis age of breast cancer and incident heart failure.

**Figure S9.** Kaplan Meier curves of the association between breast cancer and incident myocardial infarction in participants with breast cancer diagnosed at age <50 and their controls (n=10 696).

**Figure S10.** Kaplan Meier curves of the association between breast cancer and incident myocardial infarction in participants with breast cancer diagnosed at age 50 to 59 and their controls (n=22 548).

**Figure S11.** Kaplan Meier curves of the association between breast cancer and incident myocardial infarction in participants with breast cancer diagnosed at age ≥60 and their controls (n=31 720).

**Figure S12.** Kaplan Meier curves of the association between breast cancer and incident heart failure in participants with breast cancer diagnosed at age <50 and their controls (n=10 696).

**Figure S13.** Kaplan Meier curves of the association between breast cancer and incident myocardial infarction in participants with heart failure diagnosed at age 50 to 59 and their controls (n=22 548).

**Figure S14.** Kaplan Meier curves of the association between breast cancer and incident heart failure in participants with breast cancer diagnosed at age ≥60 and their controls (n=31 720).

**Table S1. Ascertainment of breast cancer and age at breast cancer diagnosis.**

|  | **Definition** | **Assessment** | **UK biobank Data-Field ID** |
| --- | --- | --- | --- |
| Breast cancer | Yes, No | (1) Cancer register: breast cancer (the International Classification of Diseases Tenth Revision [ICD-10] codes of C50). | 40006 |
| Breast cancer diagnosis age | Years | (1) Cancer register: age at cancer diagnosis (the International Classification of Diseases Tenth Revision [ICD-10] codes of C50). | 40008 |

**Table S2. Ascertainment of myocardial infarction and heart failure.**

| **Outcomes** | **Definition** | **Assessment** | **UK biobank Data-Field ID** |
| --- | --- | --- | --- |
| Myocardial infarction | Yes, No | (1) Algorithmically-defined outcomes: date of myocardial infarction, STEMI, and NSTEMI. | 42000, 42002, 42004 |
| Heart failure | Yes, No | (1) First occurrences of circulatory system disorders: heart failure (the International Classification of Diseases Tenth Revision [ICD-10] codes of I50). | 131354 |

**Table S3. Definition and assessment of covariates.**

| **Covariates** | **Definition** | **Assessment** | **UK biobank Data-Field ID** |
| --- | --- | --- | --- |
| Age (years) | Age in years | Difference between date attended baseline assessment and date of birth recorded by NHS | 21003 |
| Ethnicity | White, Nonwhite (Mixed, Asian, Black, Chinese, Other) | Touchscreen questionnaire: “What is your ethnic group?” | 21000 |
| Education | Higher education (college or university degree, other professional qualifications), other than higher education | Touchscreen questionnaire: “Which of the following qualifications do you have?” | 6138 |
| Current smoking | Yes, No | Touchscreen questionnaire: “Do you smoke tobacco now?” and “In the past, how often have you smoked tobacco?” | 20116 |
| Current drinking | At least once per week, less than once per week | Touchscreen questionnaire: “About how often do you drink alcohol?” | 1558 |
| Obesity | Yes, No | Physical measures: body mass index (BMI) ≥30 kg/m^2^ | 23104 |
| Exercise | Attending moderate or vigorous physical activity 10+ minutes at least twice per week, less than twice per week | Touchscreen questionnaire: “In a typical WEEK, on how many days did you do 10 minutes or more of moderate physical activities like carrying light loads, cycling at normal pace? (Do not include walking);  In a typical WEEK, how many days did you do 10 minutes or more of vigorous physical activity? (These are activities that make you sweat or breathe hard such as fast cycling, aerobics, heavy lifting)” | 884, 904 |
| Low-density lipoprotein cholesterol | mmol/L | Blood biochemistry: Low-density lipoprotein cholesterol | 30780 |
| Depressed mood | Yes (nearly every day or more than half the days), No (not at all or several days) | Touchscreen questionnaire: “Over the past two weeks, how often have you felt down, depressed or hopeless?” | 2050 |
| Hypertension | Yes, No | Touchscreen questionnaire and verbal interview: self-reported hypertension or antihypertensive drug use;  Average SBP/DBP ≥ 140/90 mmHg at baseline | 6150, 20002, 6177, 4079, 4080, 93, 94 |
| Diabetes | Yes, No | Touchscreen questionnaire and verbal interview: self-reported diabetes (diabetes, type 1 diabetes, or type 2 diabetes) or antidiabetic drug use;  Plasma HbA_1c_ ≥ 48 mmol/mol (6.5%) | 2443, 20002, 6153, 6177, 30750, 20003 |
| Statin use | Yes, No | Verbal interview: self-reported statin use | 20003 |
| Antihypertensive drug use | Yes, No | Touchscreen questionnaire and verbal interview: self-reported antihypertensive drug use | 6153, 6177, 20003 |
| Antidiabetic drug use | Yes, No | Touchscreen questionnaire and verbal interview: self-reported antidiabetic drug use | 6153, 6177, 20003 |

**Table S4. Baseline characteristics of participants by breast cancer status after propensity score matching (n=64 964).**

| **Characteristic** | **Breast cancer** | **Non-breast cancer** | **Effect size**^a^ |
| --- | --- | --- | --- |
| **< 50 years (n=10 696, median follow-up=13 years, interquartile range: 12-13 years)** | | | |
| Age, years | 52.1±7.4 | 52.1±7.4 | -0.001 |
| White | 2 514 (94.0) | 7 563 (94.3) | -0.005 |
| Higher education | 1 382 (51.7) | 4 139 (51.6) | 0.001 |
| Current smoking | 257 (9.6) | 756 (9.4) | 0.003 |
| Current drinking | 1 727 (64.6) | 5 303 (66.1) | -0.014 |
| Obesity | 505 (18.9) | 1 507 (18.8) | 0.001 |
| Exercise | 2 048 (76.6) | 6 132 (76.4) | 0.002 |
| LDL-C, mmol/L | 3.53±0.88 | 3.52±0.84 | 0.022 |
| Depressed mood | 174 (6.5) | 498 (6.2) | 0.005 |
| Hypertension | 976 (36.5) | 2 938 (36.6) | -0.001 |
| Diabetes | 90 (3.4) | 250 (3.1) | 0.006 |
| Antihypertensive drug use | 272 (10.2) | 795 (9.9) | 0.004 |
| Antidiabetic drug use | 54 (2.0) | 154 (1.9) | 0.003 |
| Statin use | 168 (6.3) | 485 (6.1) | 0.004 |
| **50-59 years (n=22 548, median follow-up=13 years, interquartile range: 12-13 years)** | | | |
| Age, years | 56.7±7.2 | 56.7±7.8 | 0.003 |
| White | 5 416 (96.1) | 16 252 (96.1) | -0.001 |
| Higher education | 2 767 (49.1) | 8 417 (49.8) | -0.006 |
| Current smoking | 530 (9.4) | 1 588 (9.4) | <0.001 |
| Current drinking | 3 650 (64.8) | 10 925 (64.6) | 0.001 |
| Obesity | 1 339 (23.8) | 3 949 (23.4) | 0.004 |
| Exercise | 4 334 (76.9) | 13 056 (77.2) | -0.003 |
| LDL-C, mmol/L | 3.64±0.88 | 3.66±0.87 | -0.020 |
| Depressed mood | 287 (5.1) | 867 (5.1) | -0.001 |
| Hypertension | 2 686 (47.7) | 7 908 (46.8) | 0.008 |
| Diabetes | 261 (4.6) | 754 (4.5) | 0.004 |
| Antihypertensive drug use | 900 (16.0) | 2 662 (15.7) | 0.003 |
| Antidiabetic drug use | 156 (2.8) | 459 (2.7) | 0.001 |
| Statin use | 536 (9.5) | 1 620 (9.6) | -0.001 |
| **≥ 60 years (n=31 720, median follow-up=13 years, interquartile range: 12-14 years)** | | | |
| Age, years | 62.7±4.7 | 62.7±4.8 | -0.004 |
| White | 7 689 (97.0) | 23 057 (96.9) | 0.001 |
| Higher education | 3 422 (43.2) | 10 349 (43.5) | -0.003 |
| Current smoking | 603 (7.6) | 1 780 (7.5) | 0.002 |
| Current drinking | 5 074 (64.0) | 15 275 (64.2) | -0.002 |
| Obesity | 2 090 (26.4) | 6 184 (26.0) | 0.004 |
| Exercise | 6 124 (77.2) | 18 493 (77.7) | -0.005 |
| LDL-C, mmol/L | 3.74±0.88 | 3.74±0.88 | -0.006 |
| Depressed mood | 317 (4.0) | 932 (3.9) | 0.002 |
| Hypertension | 4 981 (62.8) | 14 791 (62.2) | 0.006 |
| Diabetes | 444 (5.6) | 1 299 (5.5) | 0.003 |
| Antihypertensive drug use | 2 031 (25.6) | 6 024 (25.3) | 0.003 |
| Antidiabetic drug use | 239 (3.0) | 722 (3.0) | -0.001 |
| Statin use | 1 181 (14.9) | 3 665 (15.4) | -0.006 |

The results are presented as the mean ± standard deviation, or No. (%).

^a^The effect sizes are standardized mean differences for continuous outcomes and the Phi coefficient for dichotomous outcomes.

LDL-C, low-density lipoprotein cholesterol.

**Table S5. Associations of age at breast cancer diagnosis with incident myocardial infarction and heart failure among participants with breast cancer: competing risk models (n=16 241).**

| **Outcome** | **HR (95% CI)**^a^ | ***P* value** |
| --- | --- | --- |
| Myocardial infarction |  |  |
| ≥ 60 years (n=7 930) | Reference | / |
| 50-59 years (n=5 637) | 1.02 (0.76 to 1.37) | 0.896 |
| < 50 years (n=2 674) | 2.11 (1.47 to 3.04) | <0.001 |
| Per 10-year decrease | 1.33 (1.16 to 1.53) | <0.001 |
| Heart failure |  |  |
| ≥ 60 years (n=7 930) | Reference | / |
| 50-59 years (n=5 637) | 1.29 (1.04 to 1.59) | 0.019 |
| < 50 years (n=2 674) | 1.62 (1.18 to 2.23) | 0.003 |
| Per 10-year decrease | 1.28 (1.15 to 1.43) | <0.001 |

^a^Adjusted for age, ethnicity, education, current smoking, current drinking, obesity, exercise, low-density lipoprotein cholesterol, depressed mood, hypertension, diabetes, antihypertensive drug use, antidiabetic drug use, and statin use.

HR, hazard ratio; CI, confidence interval.

**Table S6. Associations of breast cancer with incident myocardial infarction and heart failure among different diagnosis age groups after propensity score matching: competing risk models (n=64 964).**

| **Outcome** | **HR (95% CI)**^a^  **Breast cancer vs. Non-breast cancer** | ***P* value** |
| --- | --- | --- |
| Myocardial infarction |  |  |
| ≥ 60 years (n=31 720) | 0.73 (0.61 to 0.87) | <0.001 |
| 50-59 years (n=22 548) | 0.71 (0.55 to 0.92) | 0.010 |
| < 50 years (n=10 696) | 1.64 (1.14 to 2.37) | 0.008 |
| Heart failure |  |  |
| ≥ 60 years (n=31 720) | 1.01 (0.88 to 1.17) | 0.885 |
| 50-59 years (n=22 548) | 1.31 (1.07 to 1.60) | 0.008 |
| < 50 years (n=10 696) | 2.09 (1.45 to 2.99) | <0.001 |

^a^Adjusted for age, ethnicity, education, current smoking, current drinking, obesity, exercise, low-density lipoprotein cholesterol, depressed mood, hypertension, diabetes, antihypertensive drug use, antidiabetic drug use, and statin use.

HR, hazard ratio; CI, confidence interval.

**Table S7. Associations of age at breast cancer diagnosis with incident myocardial infarction and heart failure among participants with breast cancer after excluding myocardial infarction and heart failure diagnosed within 5 years since baseline (n=15 589).**

| **Outcome** | **HR (95% CI)**^a^ | ***P* value** |
| --- | --- | --- |
| Myocardial infarction |  |  |
| ≥ 60 years (n=7 722) | Reference | / |
| 50-59 years (n=5 368) | 0.80 (0.58 to 1.12) | 0.196 |
| < 50 years (n=2 499) | 1.58 (1.03 to 2.43) | 0.038 |
| Per 10-year decrease | 1.13 (0.97 to 1.32) | 0.131 |
| Heart failure |  |  |
| ≥ 60 years (n=7 722) | Reference | / |
| 50-59 years (n=5 368) | 1.24 (0.99 to 1.56) | 0.066 |
| < 50 years (n=2 499) | 1.45 (1.00 to 2.09) | 0.049 |
| Per 10-year decrease | 1.21 (1.07 to 1.37) | 0.002 |

^a^Adjusted for age, ethnicity, education, current smoking, current drinking, obesity, exercise, low-density lipoprotein cholesterol, depressed mood, hypertension, diabetes, antihypertensive drug use, antidiabetic drug use, and statin use.

HR, hazard ratio; CI, confidence interval.

**Table S8. Associations of breast cancer with incident myocardial infarction and heart failure among different diagnosis age groups after excluding myocardial infarction and heart failure diagnosed within 5 years since baseline, results from propensity score matching analyses (n=62 356).**

| **Outcome** | **HR (95% CI)**^a^  **Breast cancer vs. Non-breast cancer** | ***P* value** |
| --- | --- | --- |
| Myocardial infarction |  |  |
| ≥ 60 years (n=30 888) | 0.86 (0.71 to 1.03) | 0.108 |
| 50-59 years (n=21 472) | 0.65 (0.48 to 0.88) | 0.005 |
| < 50 years (n=9 996) | 1.16 (0.76 to 1.77) | 0.494 |
| Heart failure |  |  |
| ≥ 60 years (n=30 888) | 1.19 (1.02 to 1.40) | 0.028 |
| 50-59 years (n=21 472) | 1.37 (1.10 to 1.71) | 0.005 |
| < 50 years (n=9 996) | 2.49 (1.61 to 3.85) | <0.001 |

^a^Adjusted for age, ethnicity, education, current smoking, current drinking, obesity, exercise, low-density lipoprotein cholesterol, depressed mood, hypertension, diabetes, antihypertensive drug use, antidiabetic drug use, and statin use.

HR, hazard ratio; CI, confidence interval.

**Table S9. Associations of age at breast cancer diagnosis with incident myocardial infarction and heart failure among participants with breast cancer after excluding participants aged <50 years at baseline (n=14 000).**

| **Outcome** | **HR (95% CI)**^a^ | ***P* value** |
| --- | --- | --- |
| Myocardial infarction |  |  |
| ≥ 60 years (n=7 907) | Reference | / |
| 50-59 years (n=4 533) | 1.08 (0.81 to 1.44) | 0.611 |
| < 50 years (n=1 560) | 2.08 (1.42 to 3.04) | <0.001 |
| Per 10-year decrease | 1.35 (1.17 to 1.55) | <0.001 |
| Heart failure |  |  |
| ≥ 60 years (n=7 907) | Reference | / |
| 50-59 years (n=4 533) | 1.32 (1.06 to 1.64) | 0.012 |
| < 50 years (n=1 560) | 1.68 (1.20 to 2.36) | 0.003 |
| Per 10-year decrease | 1.31 (1.18 to 1.46) | <0.001 |

^a^Adjusted for age, ethnicity, education, current smoking, current drinking, obesity, exercise, low-density lipoprotein cholesterol, depressed mood, hypertension, diabetes, antihypertensive drug use, antidiabetic drug use, and statin use.

HR, hazard ratio; CI, confidence interval.

**Table S10. Associations of breast cancer with incident myocardial infarction and heart failure among different diagnosis age groups after excluding participants aged <50 years at baseline, results from propensity score matching analyses (n=56 000).**

| **Outcome** | **HR (95% CI)**^a^  **Breast cancer vs. Non-breast cancer** | ***P* value** |
| --- | --- | --- |
| Myocardial infarction |  |  |
| ≥ 60 years (n=31 628) | 0.80 (0.68 to 0.96) | 0.013 |
| 50-59 years (n=18 132) | 0.84 (0.65 to 1.10) | 0.201 |
| < 50 years (n=6 240) | 1.60 (1.07 to 2.39) | 0.022 |
| Heart failure |  |  |
| ≥ 60 years (n=31 628) | 1.04 (0.90 to 1.20) | 0.627 |
| 50-59 years (n=18 132) | 1.30 (1.06 to 1.60) | 0.013 |
| < 50 years (n=6 240) | 1.58 (1.08 to 2.30) | 0.018 |

^a^Adjusted for age, ethnicity, education, current smoking, current drinking, obesity, exercise, low-density lipoprotein cholesterol, depressed mood, hypertension, diabetes, antihypertensive drug use, antidiabetic drug use, and statin use.

HR, hazard ratio; CI, confidence interval.

**Table S11. Associations of age at breast cancer diagnosis with incident myocardial infarction and heart failure among participants with breast cancer when the follow-up period ends on December 31, 2019 (n=15 909).**

| **Outcome** | **HR (95% CI)**^a^ | ***P* value** |
| --- | --- | --- |
| Myocardial infarction |  |  |
| ≥ 60 years (n=7 672) | Reference | / |
| 50-59 years (n=5 563) | 1.31 (0.94 to 1.82) | 0.109 |
| < 50 years (n=2 674) | 2.52 (1.66 to 3.84) | <0.001 |
| Per 10-year decrease | 1.51 (1.29 to 1.76) | <0.001 |
| Heart failure |  |  |
| ≥ 60 years (n=7 672) | Reference | / |
| 50-59 years (n=5 563) | 1.56 (1.22 to 1.99) | <0.001 |
| < 50 years (n=2 674) | 1.77 (1.22 to 2.58) | 0.003 |
| Per 10-year decrease | 1.38 (1.22 to 1.57) | <0.001 |

^a^Adjusted for age, ethnicity, education, current smoking, current drinking, obesity, exercise, low-density lipoprotein cholesterol, depressed mood, hypertension, diabetes, antihypertensive drug use, antidiabetic drug use, and statin use.

HR, hazard ratio; CI, confidence interval.

**Table S12. Associations of breast cancer with incident myocardial infarction and heart failure among different diagnosis age groups when the follow-up period ends on December 31, 2019, results from propensity score matching analyses (n=63 636).**

| **Outcome** | **HR (95% CI)**^a^  **Breast cancer vs. Non-breast cancer** | ***P* value** |
| --- | --- | --- |
| Myocardial infarction |  |  |
| ≥ 60 years (n=30 688) | 0.62 (0.50 to 0.76) | <0.001 |
| 50-59 years (n=22 252) | 0.82 (0.61 to 1.08) | 0.158 |
| < 50 years (n=10 696) | 1.21 (0.81 to 1.79) | 0.352 |
| Heart failure |  |  |
| ≥ 60 years (n=30 688) | 0.86 (0.73 to 1.02) | 0.088 |
| 50-59 years (n=22 252) | 1.57 (1.25 to 1.96) | <0.001 |
| < 50 years (n=10 696) | 1.37 (0.93 to 2.02) | 0.115 |

^a^Adjusted for age, ethnicity, education, current smoking, current drinking, obesity, exercise, low-density lipoprotein cholesterol, depressed mood, hypertension, diabetes, antihypertensive drug use, antidiabetic drug use, and statin use.

HR, hazard ratio; CI, confidence interval.

**Table S13. Associations of age at breast cancer diagnosis with incident myocardial infarction and heart failure among participants with breast cancer after further adjusting for menopausal status, breast cancer surgery, and hormone replacement therapy (n=16 241).**

| **Outcome** | **HR (95% CI)**^a^ | ***P* value** | **Number (%)** | | |
| --- | --- | --- | --- | --- | --- |
|  |  |  | **Menopause** | **Surgery** | **HRT** |
| Myocardial infarction |  |  |  |  |  |
| ≥ 60 years (n=7 672) | Reference | / | 6 503 (82.0%) | 7 453 (94.0%) | 4 362 (55.0%) |
| 50-59 years (n=5 563) | 1.05 (0.78 to 1.41) | 0.744 | 3 535 (62.7%) | 4 789 (85.0%) | 2 134 (37.9%) |
| < 50 years (n=2 674) | 2.19 (1.53 to 3.16) | <0.001 | 1 422 (53.2%) | 1 983 (74.2%) | 288 (10.8 %) |
| Per 10-year decrease | 1.36 (1.19 to 1.56) | <0.001 | 11 460 (70.6 %) | 14 225 (87.6%) | 6 784 (41.8%) |
| Heart failure |  |  |  |  |  |
| ≥ 60 years (n=7672) | Reference | / | 6 503 (82.0%) | 7 453 (94.0%) | 4 362 (55.0%) |
| 50-59 years (n=5563) | 1.33 (1.08 to 1.64) | 0.008 | 3 535 (62.7%) | 4 789 (85.0%) | 2 134 (37.9%) |
| < 50 years (n=2674) | 1.74 (1.25 to 2.41) | 0.001 | 1 422 (53.2%) | 1 983 (74.2%) | 288 (10.8 %) |
| Per 10-year decrease | 1.33 (1.19 to 1.48) | <0.001 | 11 460 (70.6 %) | 14 225 (87.6%) | 6 784 (41.8%) |

^a^Adjusted for age, ethnicity, education, current smoking, current drinking, obesity, exercise, low-density lipoprotein cholesterol, depressed mood, hypertension, diabetes, antihypertensive drug use, antidiabetic drug use, statin use, menopausal status, breast cancer surgery, and hormone replacement therapy.

HR, hazard ratio; CI, confidence interval.

**Table S14. Associations of breast cancer with incident myocardial infarction and heart failure among different diagnosis age groups after further adjusting for menopausal status, breast cancer surgery, and hormone replacement therapy, results from propensity score matching analyses (n=64 964).**

| **Outcome** | **HR (95% CI)**^a^  **Breast cancer vs. Non-breast cancer** | ***P* value** |
| --- | --- | --- |
| Myocardial infarction |  |  |
| ≥ 60 years (n=30 688) | 0.75 (0.63 to 0.89) | 0.001 |
| 50-59 years (n=22 252) | 0.75 (0.58 to 0.98) | 0.032 |
| < 50 years (n=10 696) | 1.47 (1.03 to 2.61) | 0.036 |
| Heart failure |  |  |
| ≥ 60 years (n=30 688) | 1.03 (0.89 to 1.19) | 0.669 |
| 50-59 years (n=22 252) | 1.39 (1.13 to 1.69) | 0.001 |
| < 50 years (n=10 696) | 2.18 (1.48 to 3.20) | <0.001 |

^a^Adjusted for age, ethnicity, education, current smoking, current drinking, obesity, exercise, low-density lipoprotein cholesterol, depressed mood, hypertension, diabetes, antihypertensive drug use, antidiabetic drug use, statin use, menopausal status, breast cancer surgery, and hormone replacement therapy.

HR, hazard ratio; CI, confidence interval.

**Table S15. Comparison of baseline characteristics between participants included (n=251 277) and excluded due to history of myocardial infarction or heart failure, without complete data on low-density lipoprotein cholesterol, or having myocardial infarction or heart failure before breast cancer at follow-ups (n=22 048).**

| **Characteristics** | **Participants included**  **(n=251 277)** | **Participants excluded**  **(n=22 048)** | **Effect size**^a^ |
| --- | --- | --- | --- |
| Age, years | 56.8±8.0 | 57.5±8.0 | -0.085 |
| White | 420 508 (94.1) | 51 535 (92.5) | 0.029 |
| Higher education | 116 933 (46.5) | 8 551 (38.8) | 0.042 |
| Current smoking | 22 154 (8.8) | 2 207 (10.0) | -0.011 |
| Current drinking | 157 130 (62.5) | 12 775 (57.9) | 0.026 |
| Obesity | 58 255 (23.2) | 6 042 (27.4) | -0.027 |
| Exercise | 195 358 (77.8) | 16 473 (74.7) | 0.020 |
| Depressed mood | 13 129 (5.2) | 1 382 (6.3) | -0.013 |
| Hypertension | 121 310 (48.3) | 11 324 (51.4) | -0.017 |
| Diabetes | 10 678 (4.3) | 1 530 (6.9) | -0.036 |
| LDL-C, mmol/L | 3.63±0.87 | 2.99±0.86 | 0.739 |
| Antihypertensive drug use | 42 970 (17.1) | 4 943 (22.4) | -0.038 |
| Antidiabetic drug use | 6 077 (2.4) | 924 (4.2) | -0.031 |
| Statin use | 26 387 (10.5) | 4 033 (18.3) | -0.068 |

The results are presented as the mean ± standard deviation, or No. (%).

^a^Calculated by using a *t* test, or Chi-square test.

LDL-C, low-density lipoprotein cholesterol.

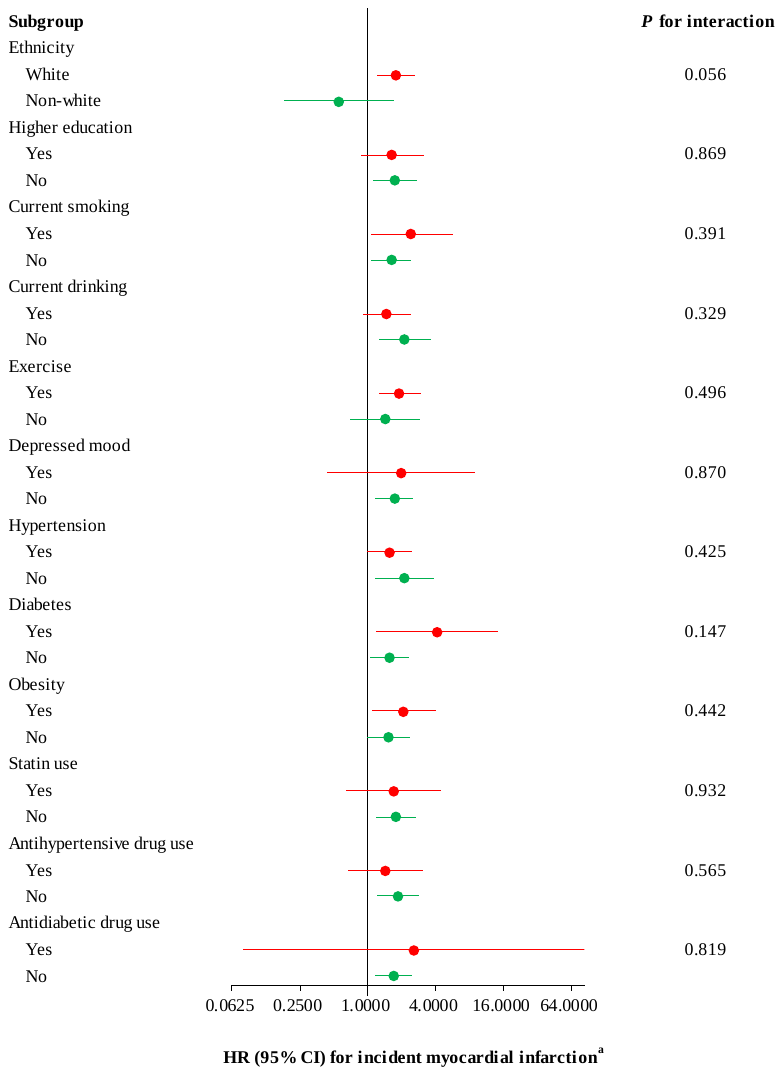

**Figure S1. Subgroup analyses to identify potential modifying effects from covariates on the associations between breast cancer and incident myocardial infarction in participants with breast cancer diagnosed at age < 50 and their controls by using Cox proportional hazards models (n=10 696).**

^a^ Adjusted for age, ethnicity, education, current smoking, current drinking, obesity, exercise, low-density lipoprotein cholesterol, depressed mood, hypertension, diabetes, antihypertensive drug use, antidiabetic drug use, and statin use, except where an adjusting variable itself was being tested, by using Cox proportional hazards models.

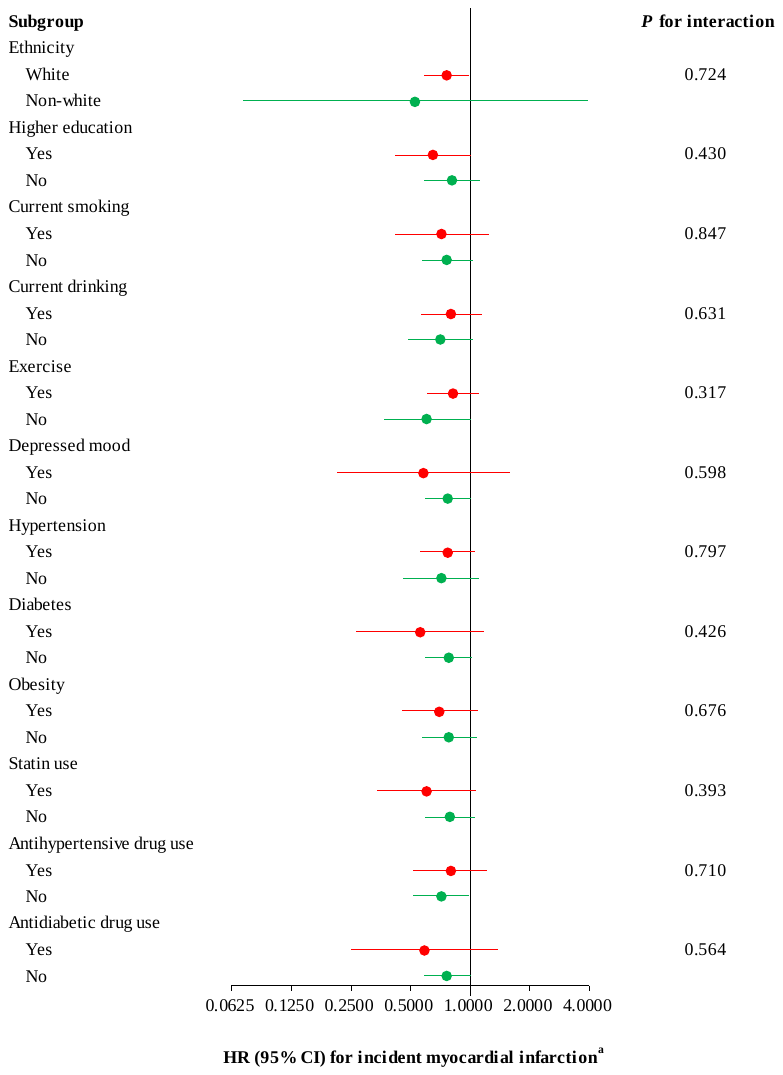

**Figure S2. Subgroup analyses to identify potential modifying effects from covariates on the associations between breast cancer and incident myocardial infarction in participants with breast cancer diagnosed at age 50 to 59 and their controls by using Cox proportional hazards models (n=22 548).**

^a^ Adjusted for age, ethnicity, education, current smoking, current drinking, obesity, exercise, low-density lipoprotein cholesterol, depressed mood, hypertension, diabetes, antihypertensive drug use, antidiabetic drug use, and statin use, except where an adjusting variable itself was being tested, by using Cox proportional hazards models.

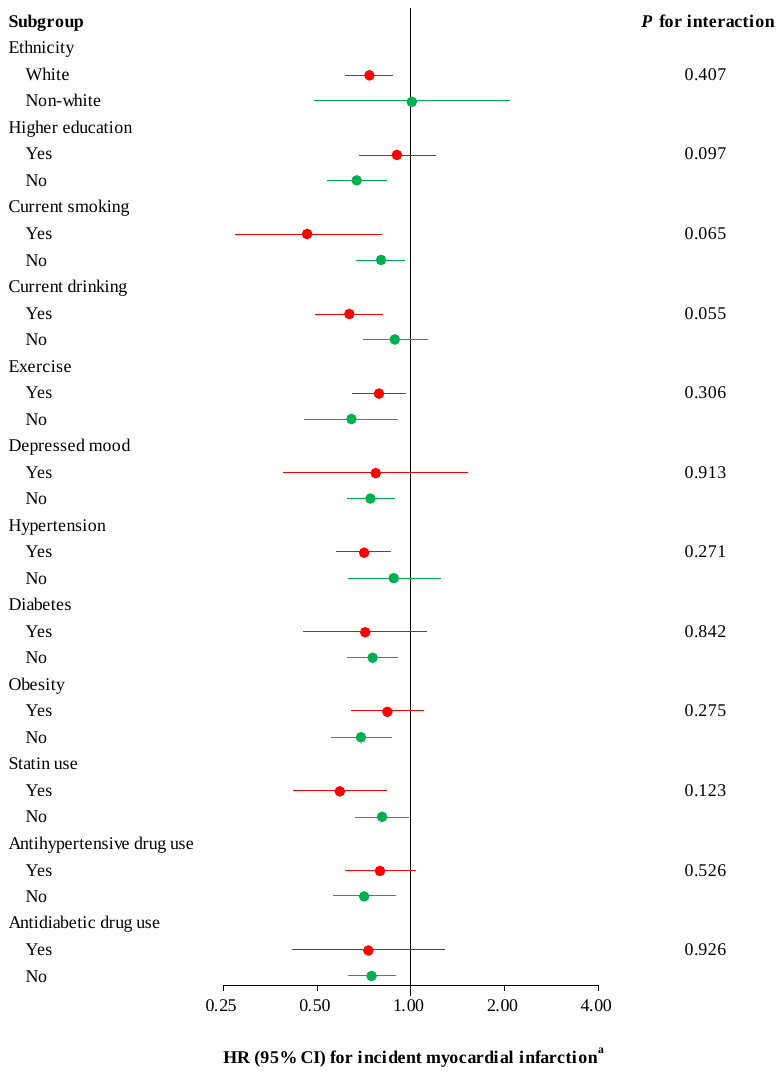

**Figure S3. Subgroup analyses to identify potential modifying effects from covariates on the associations between breast cancer and incident myocardial infarction in participants with breast cancer diagnosed at age ≥ 60 and their controls by using Cox proportional hazards models (n=31 720).**

^a^ Adjusted for age, ethnicity, education, current smoking, current drinking, obesity, exercise, low-density lipoprotein cholesterol, depressed mood, hypertension, diabetes, antihypertensive drug use, antidiabetic drug use, and statin use, except where an adjusting variable itself was being tested, by using Cox proportional hazards models.

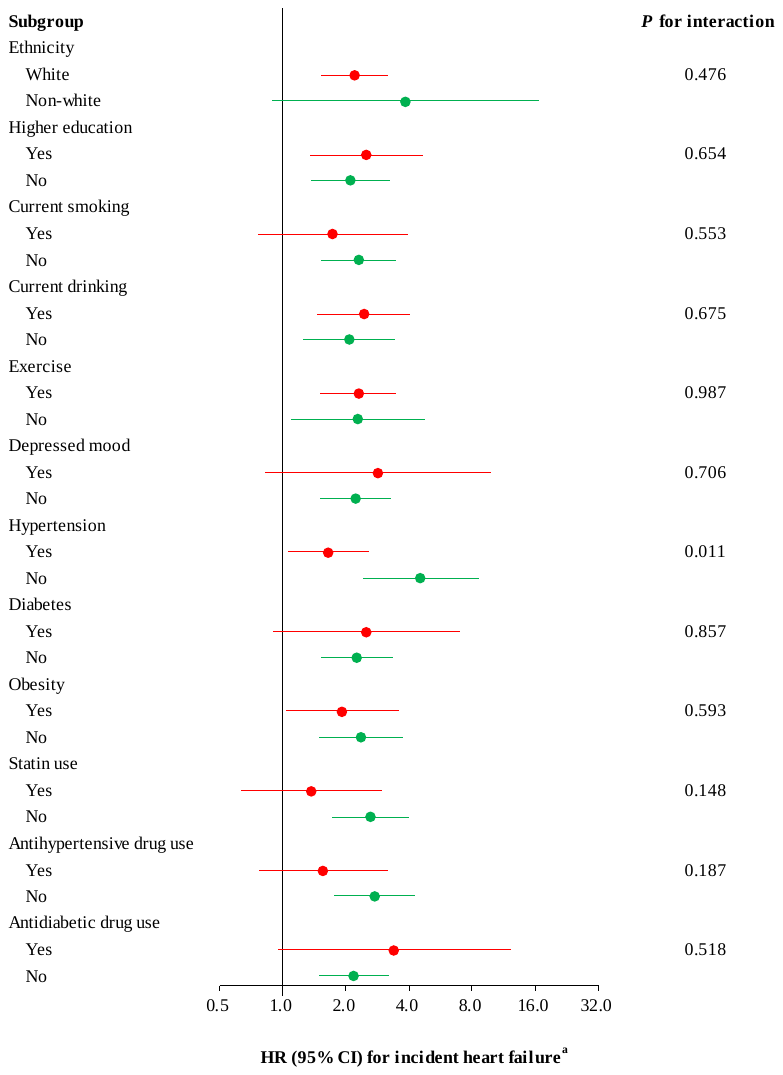

**Figure S4. Subgroup analyses to identify potential modifying effects from covariates on the associations between breast cancer and incident heart failure in participants with breast cancer diagnosed at age < 50 and their controls by using Cox proportional hazards models (n=10 696).**

^a^ Adjusted for age, ethnicity, education, current smoking, current drinking, obesity, exercise, low-density lipoprotein cholesterol, depressed mood, hypertension, diabetes, antihypertensive drug use, antidiabetic drug use, and statin use, except where an adjusting variable itself was being tested, by using Cox proportional hazards models.

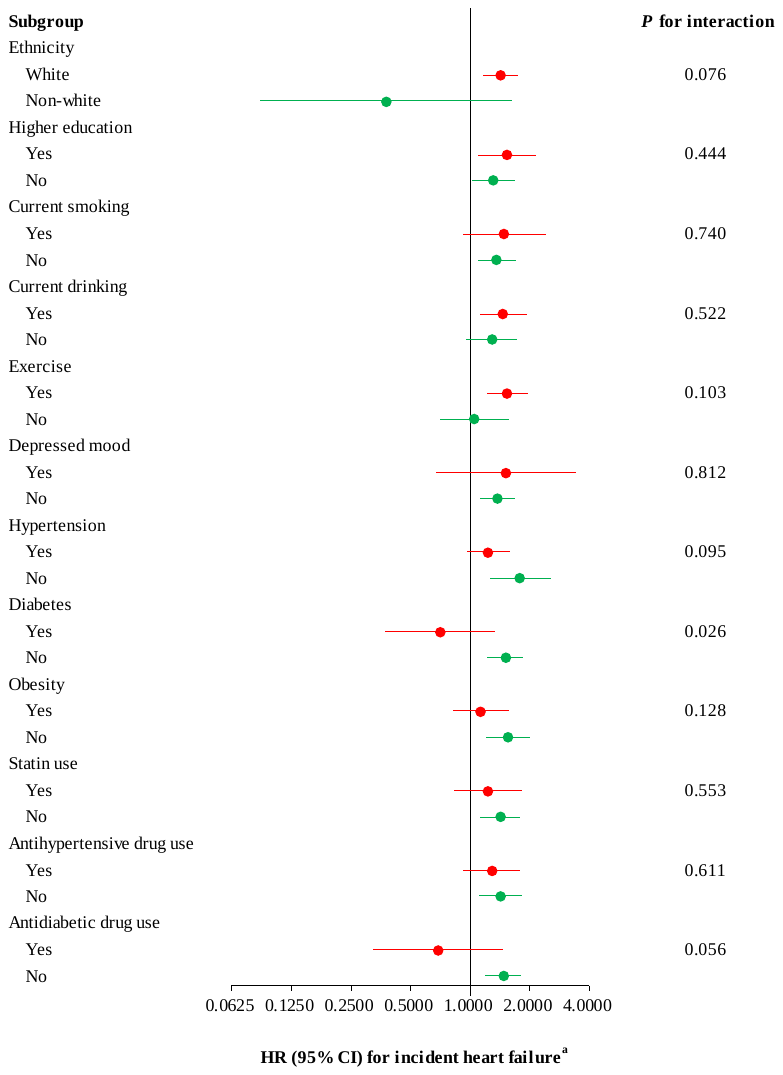

**Figure S5. Subgroup analyses to identify potential modifying effects from covariates on the associations between breast cancer and incident heart failure in participants with breast cancer diagnosed at age 50 to 59 and their controls by using Cox proportional hazards models (n=22 548).**

^a^ Adjusted for age, ethnicity, education, current smoking, current drinking, obesity, exercise, low-density lipoprotein cholesterol, depressed mood, hypertension, diabetes, antihypertensive drug use, antidiabetic drug use, and statin use, except where an adjusting variable itself was being tested, by using Cox proportional hazards models.

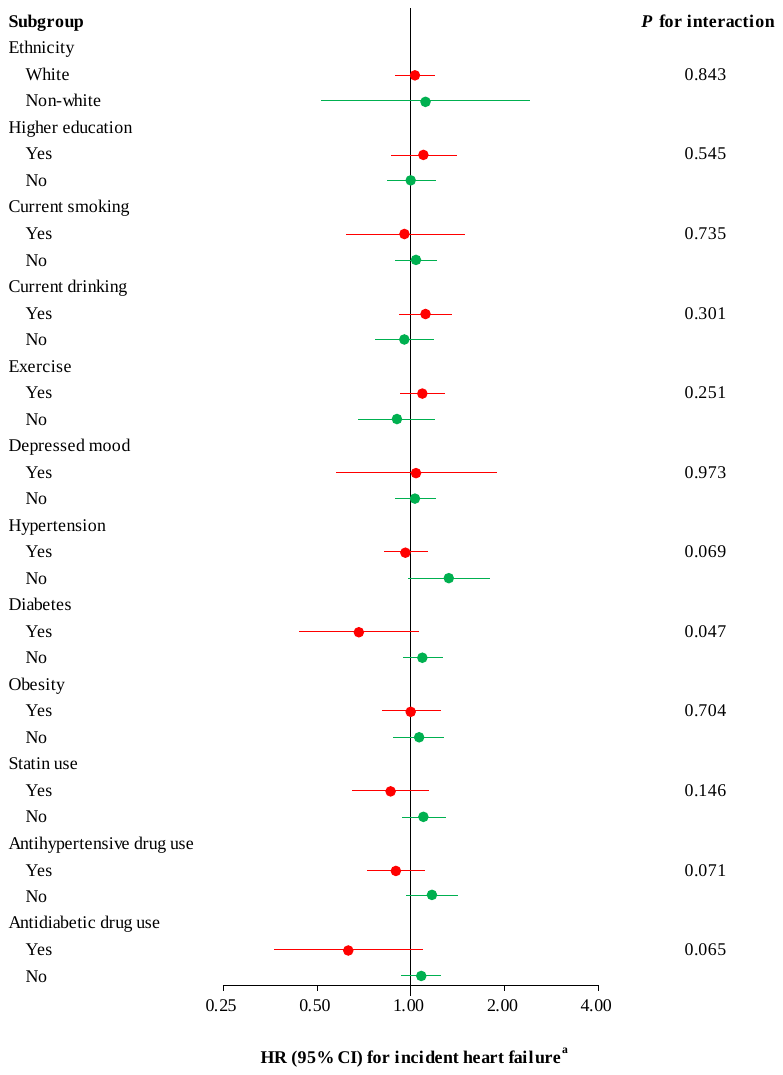

**Figure S6. Subgroup analyses to identify potential modifying effects from covariates on the associations between breast cancer and incident heart failure in participants with breast cancer diagnosed at age ≥ 60 and their controls by using Cox proportional hazards models (n=31 720).**

^a^ Adjusted for age, ethnicity, education, current smoking, current drinking, obesity, exercise, low-density lipoprotein cholesterol, depressed mood, hypertension, diabetes, antihypertensive drug use, antidiabetic drug use, and statin use, except where an adjusting variable itself was being tested, by using Cox proportional hazards models.

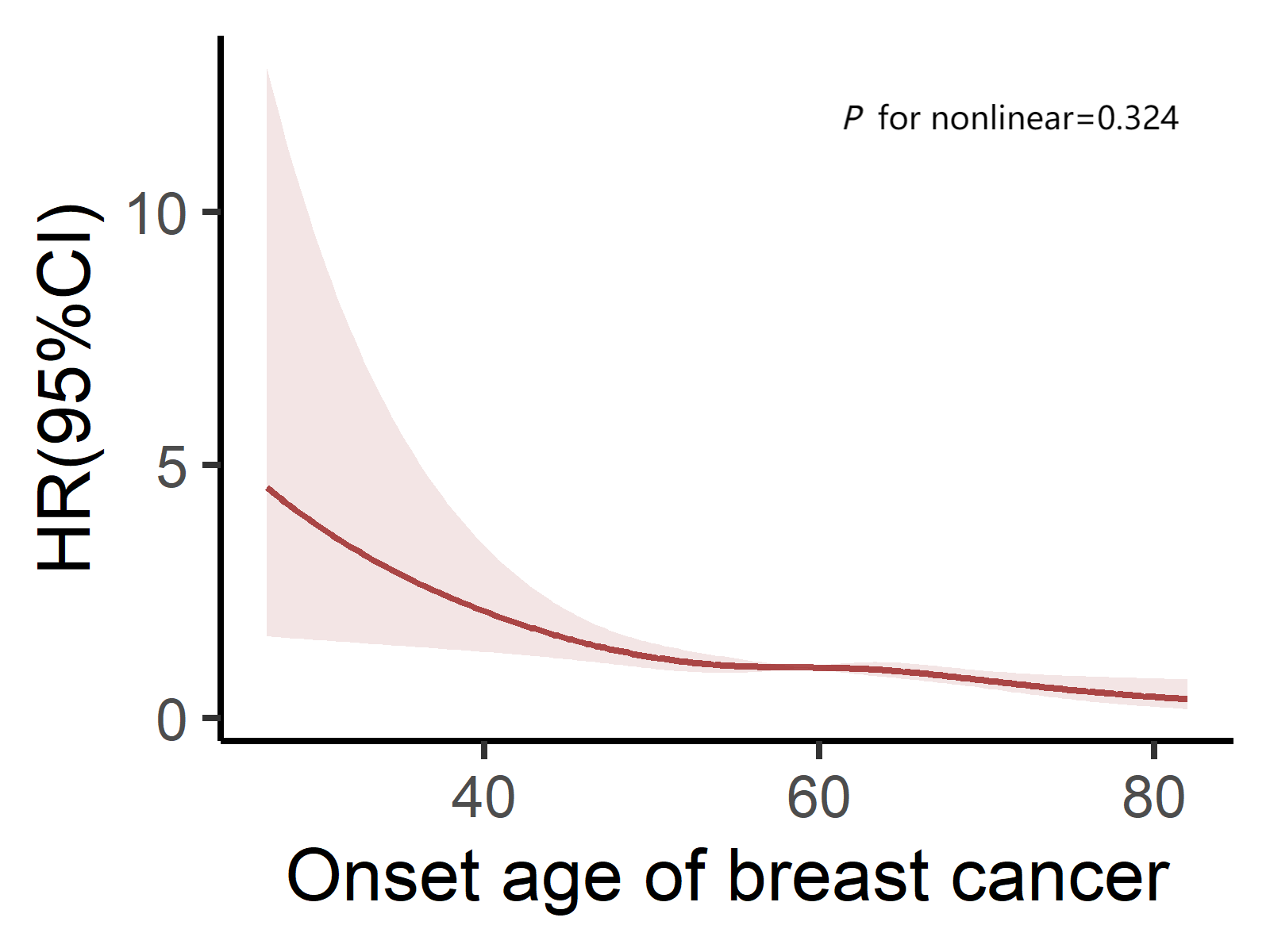

**Figure S7. Cubic spline curves of the association between diagnosis age of breast cancer and incident myocardial infarction.**

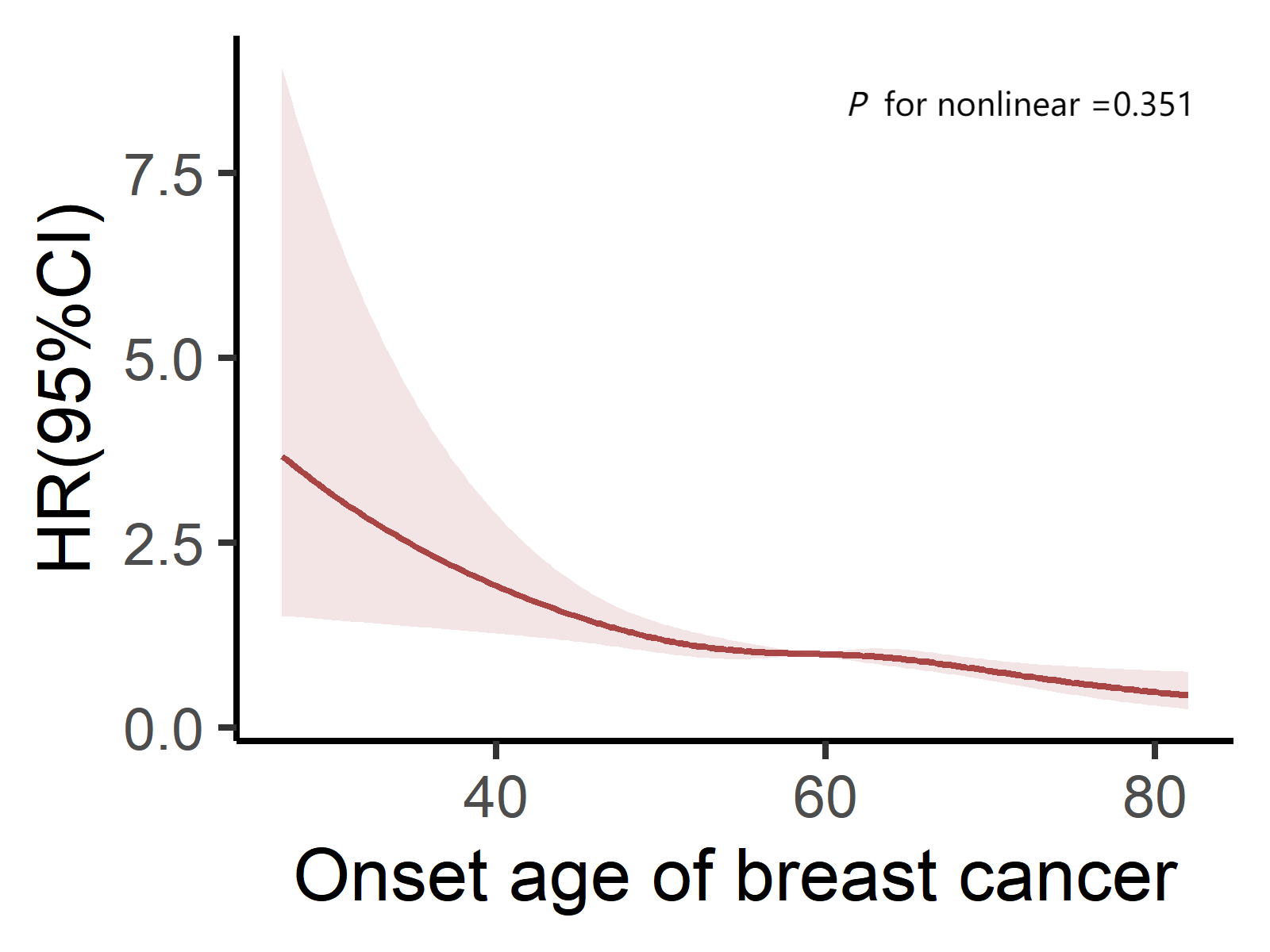

**Figure S8. Cubic spline curves of the association between diagnosis age of breast cancer and incident heart failure.**

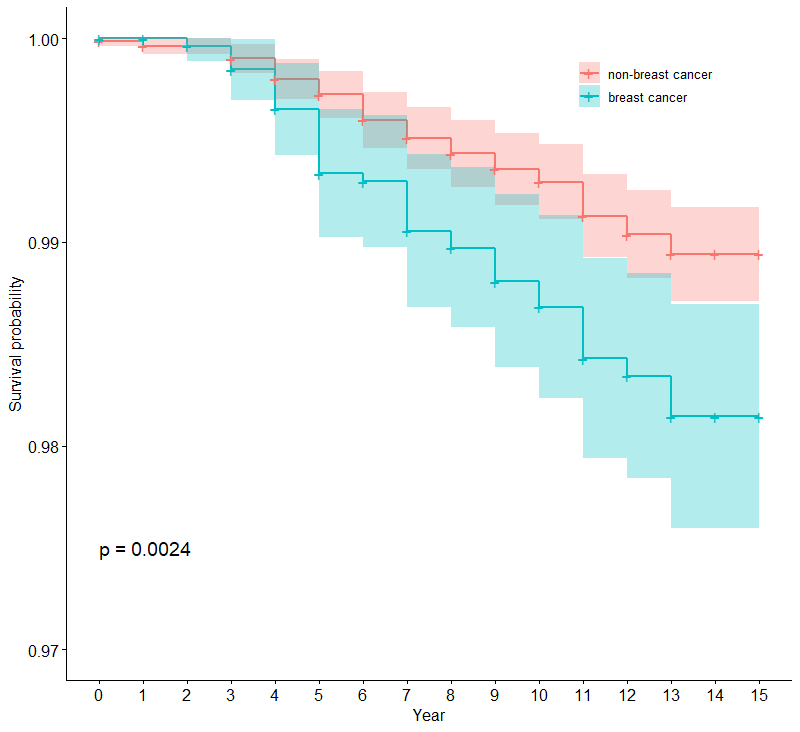
**Figure S9. Kaplan Meier curves of the association between breast cancer and incident myocardial infarction in participants with breast cancer diagnosed at age <50 and their controls (n=10 696).**

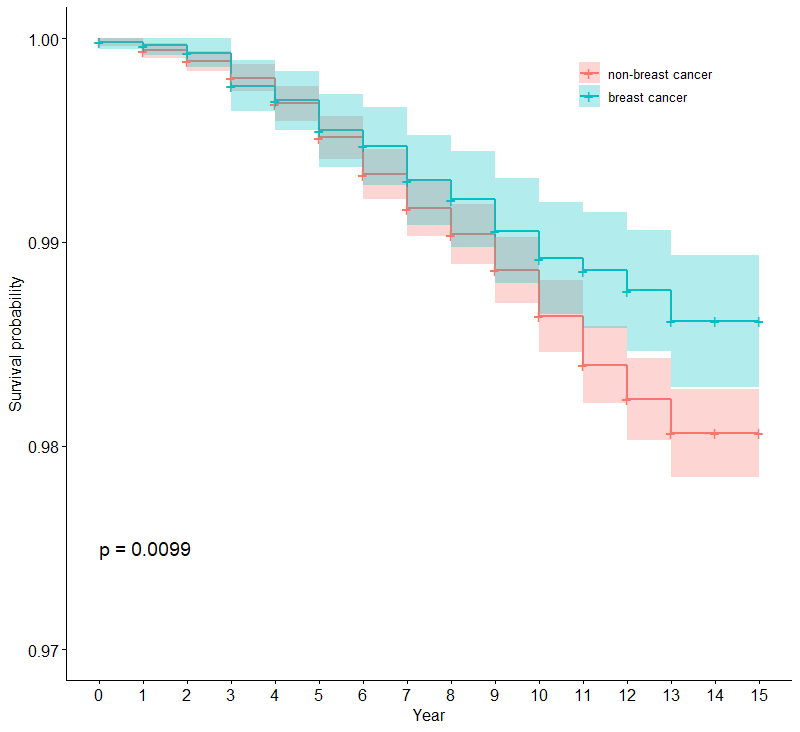
**Figure S10. Kaplan Meier curves of the association between breast cancer and incident myocardial infarction in participants with breast cancer diagnosed at age 50 to 59 and their controls (n=22 548).**

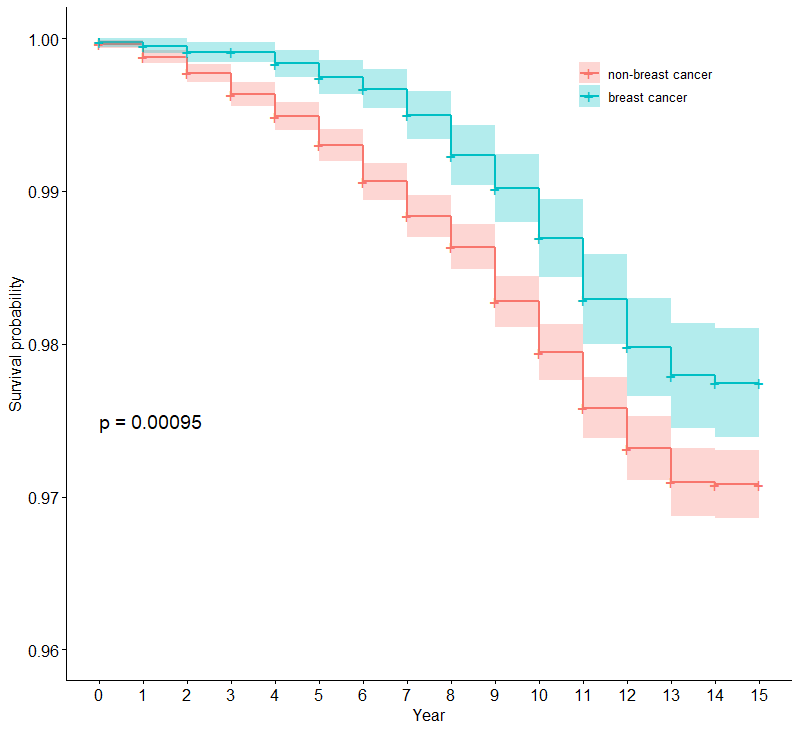
**Figure S11. Kaplan Meier curves of the association between breast cancer and incident myocardial infarction in participants with breast cancer diagnosed at age ≥60 and their controls (n=31 720).**

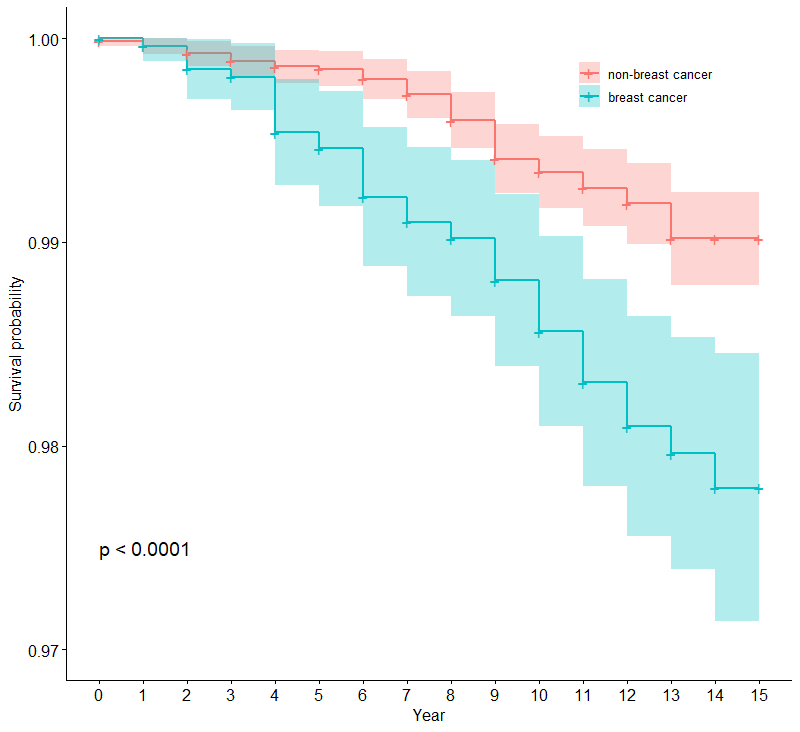
**Figure S12. Kaplan Meier curves of the association between breast cancer and incident heart failure in participants with breast cancer diagnosed at age <50 and their controls (n=10 696).**

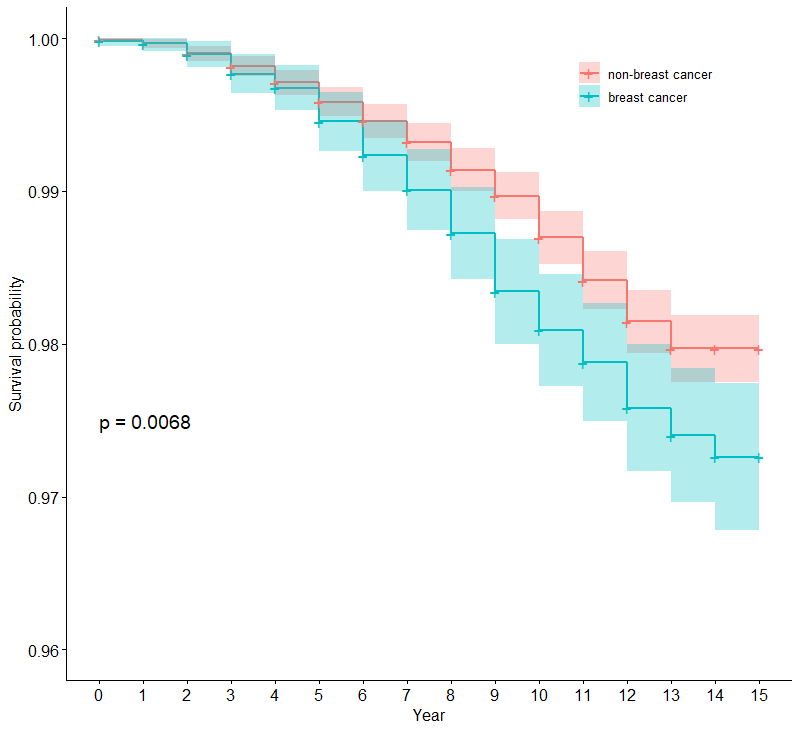
**Figure S13. Kaplan Meier curves of the association between breast cancer and incident myocardial infarction in participants with heart failure diagnosed at age 50 to 59 and their controls (n=22 548).**

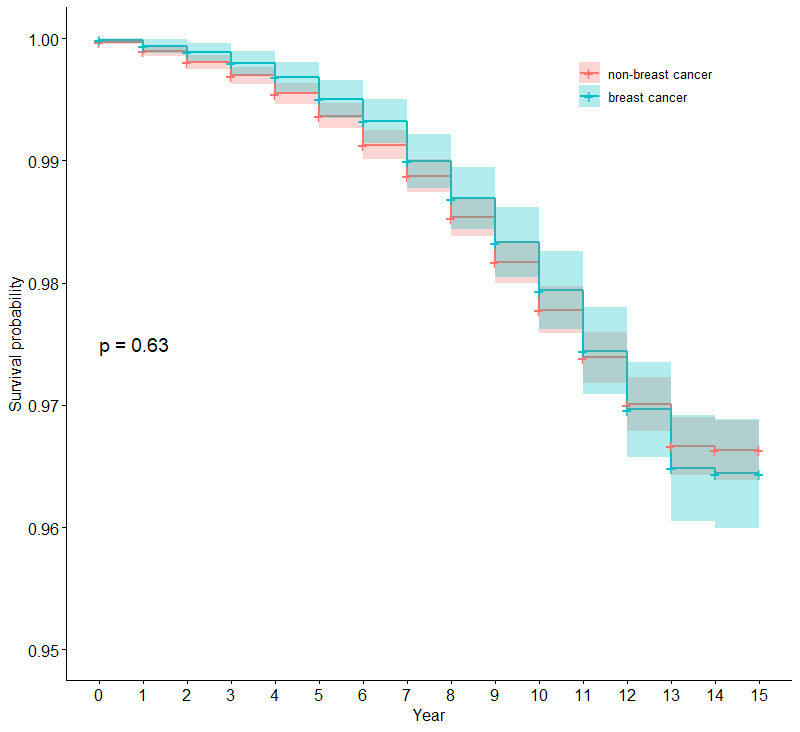
**Figure S14. Kaplan Meier curves of the association between breast cancer and incident heart failure in participants with breast cancer diagnosed at age ≥60 and their controls (n=31 720).**
